# Loneliness among older adults in the community during COVID-19

**DOI:** 10.1101/2020.11.23.20237289

**Authors:** Rachel D. Savage, Wei Wu, Joyce Li, Andrea Lawson, Susan E. Bronskill, Stephanie A. Chamberlain, Jim Grieve, Andrea Gruneir, Christina Reppas-Rindlisbacher, Nathan M. Stall, Paula A. Rochon

**Affiliations:** Women’s College Research Institute, Women’s College Hospital, Toronto, ON; ICES, Toronto, ON; Institute of Health Policy, Management and Evaluation, Dalla Lana School of Public Health, University of Toronto, Toronto, ON; Department of Family Medicine, University of Alberta, Edmonton, AB; RTOERO, Toronto, ON; Division of Geriatric Medicine, Department of Medicine, University of Toronto, Toronto, ON

## Abstract

**Objective:** Physical distancing and stay-at-home measures implemented to slow transmission of novel coronavirus disease (COVID-19) may intensify feelings of loneliness in older adults, especially those living alone. Our aim was to characterize the extent of loneliness in a sample of older adults living in the community and assess characteristics associated with loneliness.

**Design:** Online cross-sectional survey between May 6 and May 19, 2020

**Setting:** Ontario, Canada

**Participants:** Convenience sample of the members of a national retired educators’ organization.

**Primary outcome measures:** Self-reported loneliness, including differences between women and men.

**Results:** 4879 respondents (71.0% women; 67.4% 65-79 years) reported that in the preceding week, 43.1% felt lonely at least some of the time, including 8.3% that felt lonely always or often. Women had increased odds of loneliness compared to men, whether living alone (adjusted Odds Ratio (aOR) 1.52 [95% Confidence Interval (CI) 1.13-2.04]) or with others (2.44 [95% CI 2.04-2.92]). Increasing age group decreased the odds of loneliness (aOR 0.69 [95% CI 0.59-0.81] 65-79 years and 0.50 [95% CI 0.39-0.65] 80+ years compared to <65 years). Living alone was associated with loneliness, with a greater association in men (aOR 4.26 [95% CI 3.15-5.76]) than women (aOR 2.65 [95% CI 2.26-3.11]). Other factors associated with loneliness included: fair or poor health (aOR 1.93 [95% CI 1.54-2.41]), being a caregiver (aOR 1.18 [95% CI 1.02-1.37]), receiving care (aOR 1.47 [95% CI 1.19-1.81]), high concern for the pandemic (aOR 1.55 [95% CI 1.31-1.84]), not experiencing positive effects of pandemic distancing measures (aOR 1.94 [95% CI 1.62-2.32]), and changes to daily routine (aOR 2.81 [95% CI 1.96-4.03]).

**Conclusions:** While many older adults reported feeling lonely during COVID-19, several characteristics – such as being female and living alone – increased the odds of loneliness. These characteristics may help identify priorities for targeting interventions to reduce loneliness.

**Strengths and limitations of this study:**

- This survey study leveraged a strong community-based partnership to obtain timely data from a large sample of older Canadians on the impacts of COVID-19.
- This study identified several characteristics that increased the odds of loneliness, which may help to identify priorities for targeted interventions to reduce loneliness.
- The data were based on a convenience sample of retired, educational staff, who are not fully representative of the Canadian population. The perspectives of vulnerable groups who may be at greater risk for loneliness (e.g. those with severe mental health illness, low income, no home internet access, etc.) are likely underrepresented in this sample.

## Background

As data emerge on how common, yet harmful, it is to be lonely, loneliness is increasingly recognized as a public health priority. In the United States, more than 40% of respondents to the nationally representative Health and Retirement Study reported feeling lonely.^1^ In Canada, 1 in 4 older women and 1 in 5 older men report feeling lonely at least some of the time.^2^ Older adults are particularly susceptible to loneliness because of aging-related events (e.g. retirement, declining health, widowhood). Women report higher rates of loneliness than men,^2,3^ possibly due to their longer life expectancy and greater likelihood of outliving their spouse, resulting in prolonged widowhood,^4,5^ their caregiver roles,^2,6,7^ lower incomes^8^, and their greater tendency to acknowledge feeling lonely.^5^ Addressing loneliness is important because of its profound impact on health and well-being, including increased risk for premature death,^9,10^ cardiovascular disease, depression, dementia and even suicide.^11-17^

The novel coronavirus pandemic (COVID-19) and accompanying physical distancing and stay-at-home measures (i.e. closure of nonessential businesses and public spaces, as well as recommendations to practice physical distancing with anyone outside the home) are expected to intensify feelings of loneliness. Previous infectious disease outbreaks and pandemics have demonstrated increases in loneliness, anxiety, and depression from quarantine-induced social isolation.^18,19^

Understanding how older adults have been impacted by COVID-19 is vital to address their needs promptly and effectively and prevent unnecessary harms as the pandemic persists. Early cross-sectional studies have examined public concerns regarding COVID-19 (e.g. becoming infected, reduced health care access) and its impact on daily life.^20,21^ While valuable, these studies were conducted prior to or on the cusp of the implementation of physical distancing and stay-at-home measures, did not report on mental health, under-represented older adults^20^, a key high-risk group, and did not explore important differences between women and men. McGinty et al recently published prevalence estimates of psychological distress and loneliness in the US; although, subgroup analyses focused on psychological distress rather than loneliness.^22^ Timely data are needed that are relevant to older women and men to inform public health responses and healthcare delivery.

We conducted an online cross-sectional survey to assess how the COVID-19 pandemic has affected older adults living in the community in Canada. Our objective was to characterize the extent of loneliness in older adults, including differences between women and men, and examine factors associated with loneliness to identify groups likely to benefit most from intervention.

## Methods

### Study design and setting

A closed, online cross-sectional survey was administered to members of the RTOERO (formerly known as the Retired Teachers of Ontario) between May 6 and May 19, 2020. At this time in Ontario, Canada, physical distancing measures had been in place for about seven weeks; daily case and death counts were in decline after peaks in late April; and outbreaks in long-term care homes were a focus of news headlines (**Figure 1** for timeline).

**Figure 1.**
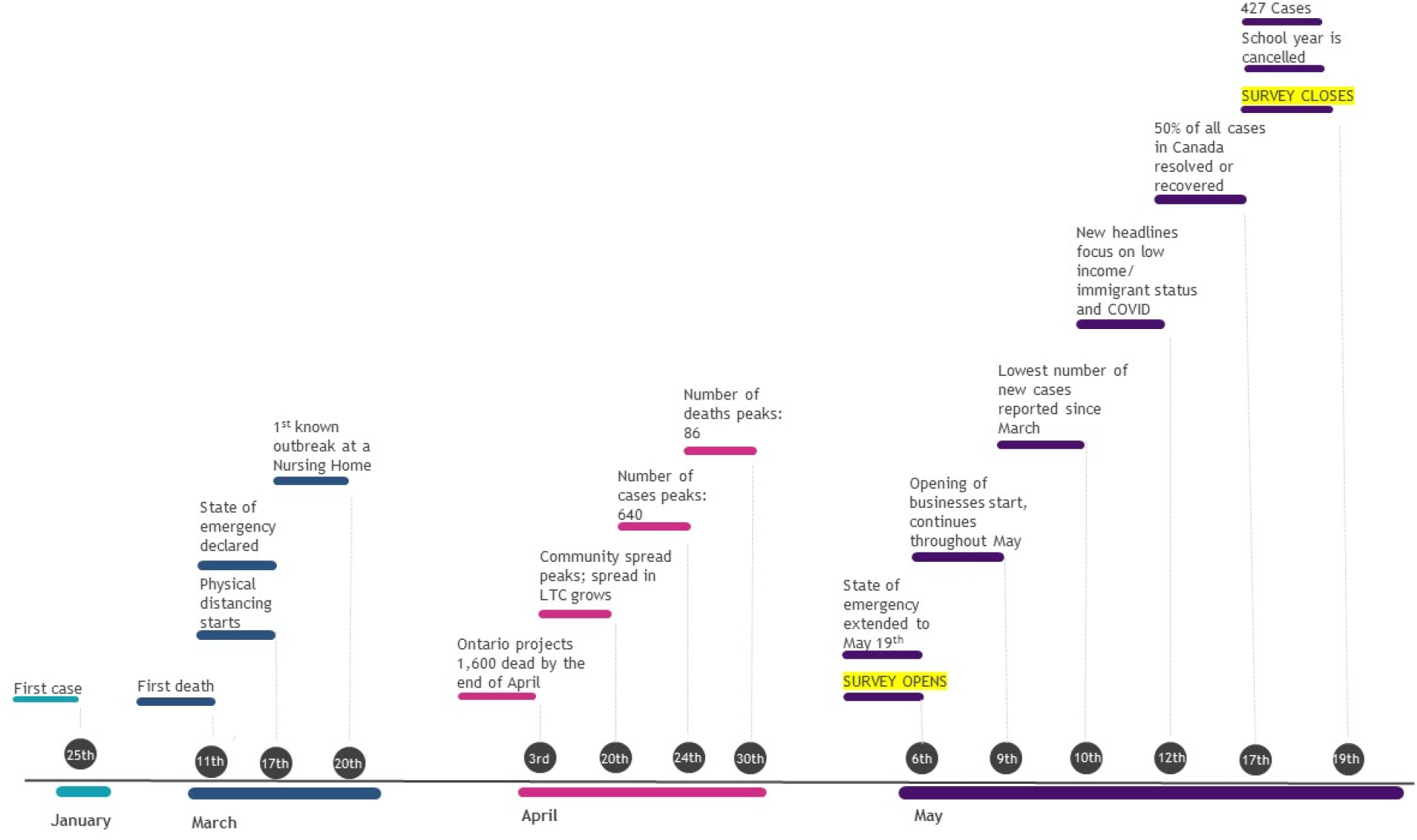
Timeline of COVID-19 in Ontario, Canada’s largest province. Physical distancing measures beginning March 17 included closure of all indoor recreational facilities, public libraries, theatres, cinemas, bars, and restaurants. Publicly funded schools were closed by this point as well, and all employers in Ontario were asked to facilitate virtual work arrangements for employees. Remaining non-essential businesses were closed March 25. Gatherings of more than 5 people were prohibited on March 28. On March 30, Ontario’s Chief Medical Officer of Health strongly recommended individuals over 70 years of age or those with compromised immune systems or underlying medical conditions to stay at home. Source: CIHI, COVID-19 Intervention Scan, Accessed Aug 11 2020, https://www.cihi.ca/en/covid-19-intervention-scan

RTOERO is a voluntary membership organization of more than 81,000 retired educators, administrators, and educational support staff, from child care, K-12 and post-secondary settings, that provides group health insurance benefits, as well as other programs and services, to the broader education community (https://www.rtoero.ca). Members were invited to participate by e-mail from RTOERO’s chief executive officer. Two reminder emails were sent at 7 and 10 days. The survey was not publicly advertised. All members were eligible to participate if they had a registered e-mail address (∼62,000). Study materials were provided in English and French. Our study design and reporting followed the Checklist for Reporting Results of Internet E-Surveys (CHERRIES).^23^

The Research Ethics Board at Women’s College Hospital in Toronto, Canada approved this study [#2020-0051-E]. A link to a study information sheet was provided on the survey’s home page and informed consent was obtained electronically. Participation was voluntary, and no incentives were provided. Minimal identifying personal information was collected (e.g. first three digits of postal code).

### Questionnaire

The questionnaire was developed with RTOERO leadership and included 32 questions (**eAppendix** in the **Supplement**). Several questions were adapted with permission from the *Stanford Coronavirus Survey* (https://pcrt.stanford.edu/covid). Questions examined the impact of COVID-19 on daily life; loneliness; and the use of digital technologies for social connectivity. We used a single-item, direct measure of loneliness by asking respondents how often they felt lonely in the past week (1-2 days, several days, most days, every day), consistent with the *Canadian Longitudinal Study on Aging* (CLSA)^2^ and the UK’s *Community Life Survey*.^24^ We chose this approach because it allowed respondents to self-report on loneliness and was considered more suitable for the pandemic context, where asking indirectly about feeling “left out” to infer loneliness may be less relevant as distancing and stay-at-home measures were universally applied.

Respondents were also asked about their history of COVID-19 symptoms and testing, the extent to which they were practising physical distancing and stay-at-home measures, and sociodemographic characteristics (i.e. age, sex, ethnicity, language, health status and location of residence). The ethnic response categories we used mirrored those used in Canada’s national health survey.^25^ The questionnaire was pretested in English with 18 RTOERO board members and staff, and in French by 1 staff member, for usability, technical functionality, clarity, flow, sensitive questions, and timing. Pretest results were not included in the final analysis.

### Patient and public involvement

As noted above, RTOERO leadership (which comprise members of RTOERO) were involved in all aspects of the study, including questionnaire development, pretesting, and participant recruitment. Preliminary results were shared with the team and feedback was incorporated into the final analysis and manuscript. RTOERO’s chief executive officer is a coauthor (JG) and critically reviewed the manuscript. Results will be shared with RTOERO members through a webinar in the fall of 2020.

### Data collection

The questionnaire was administered using SimpleSurvey™. Data were stored in an encrypted, password protected form on the secure Simple Survey server and were downloaded to the secure, password-protected Women’s College Hospital server accessible to authorized team members. All questions were optional, so completeness checks were not performed; although, respondents were reminded of unanswered questions before proceeding to the next section to minimize incomplete data. We used adaptive questioning to reduce the complexity of questions.^23,26^ Respondents were able to save their responses and return to the survey later to complete it. The survey completion rate was the number of respondents who finished the survey divided by the number consenting to participate.^23^ Surveys were only analysed if the respondent clicked “Submit” and responded to more than one question.

### Exposures

Sociodemographic characteristics - sex, age, living alone, ethnicity, rural residence, health status, and caregiver status – were collected, based on factors previously reported to be associated with loneliness.^3,4^ We additionally collected self-reported measures of social support – communication frequency, receiving offers of assistance and social media use – as well as attitudes and behaviours towards COVID-19 hypothesized to contribute to loneliness, including level of concern, change in daily routine, extent of physical distancing, and perceived positive effects of distancing measures. Variable definitions are presented in the **eMethods** in the **Supplement**.

### Outcome

Respondents were classified as lonely if they reported feeling lonely on 1 or more days in the preceding 7 days.^2,24^

### Analysis

Chi-squared tests were used to identify sex differences. To identify predictors of loneliness for older women and men, exploratory analyses using sex-stratified and sex-pooled multivariable logistic regression models were conducted. We hypothesized that loneliness would be common, particularly in women and those living alone, and that higher pandemic concern would increase loneliness. In the sex-stratified regression analysis, we calculated unadjusted and minimally adjusted (age and health status) models. In the sex-pooled model, we additionally adjusted for all covariates and formally tested for sex interactions with explanatory factors, including age group, living alone, communication frequency, receiving offers of assistance, change in daily routine, and perceived positive effects of distancing measures, identified in the stratified analysis using interaction terms. Statistical tests were two sided, with P < .05 interpreted as statistically significant. Analyses were performed using SAS version 9.4.

## Results

Overall, 5556 RTOERO members responded to the survey, of which 5509 provided consent. 4891 surveys were submitted, for a completion rate of 88.8%. We excluded 12 respondents who responded to ≤1 survey question, leaving 4879respondents included in the analysis.

### Characteristics

Most respondents were women (3421/4818 [71.0%]), between the ages of 65-79 years (3279/4863 [67.4%]) and completed the survey in English (97.6%) (**Table 1**). They were similar to the broader RTOERO membership in terms of sex (67% female), age distribution (14.5% <65 years; 64% 65-79 years; 21.5% ≥80 years) and preferred language (95% English) (personal communication, J. Grieve). One third of female respondents lived alone (1138/3356 [33.9%]) compared to one fifth of men (266/1351[19.7%]). Respondents were predominantly white (4454/4861 [91.6%]) and in good self-reported health (4370/4873 [89.7%]).

**Table 1.** Sociodemographic characteristics of older female and male survey respondents.

| **Characteristics** | **All**<br>**(N=4,879)<sup>a</sup>** | **Women**<br>**(n=3,421)** | **Men**<br>**(n=1,397)** |
| --- | --- | --- | --- |
| Language of Survey |  |  |  |
| English | 4762 (97.6%) | 3339 (97.6%) | 1365 (97.7%) |
| French | 117 (2.4%) | 82 (2.4%) | 32 (2.3%) |
| Age, years | n=4,863 | n=3,416 | n=1,395 |
| <65 | 1027 (21.1%) | 846 (24.8%) | 174 (12.5%) |
| 65-79 | 3279 (67.4%) | 2295 (67.2%) | 945 (67.7%) |
| 80+ | 557 (11.5%) | 275 (8.1%) | 276 (19.8%) |
| Living arrangement | n=4,762 | n=3,356 | n=1,351 |
| Lives alone | 1415 (29.7%) | 1138 (33.9%) | 266 (19.7%) |
| Access to private outdoor space | n=4,854 | n=3,407 | n=1,391 |
| Yes | 4706 (97.0%) | 3302 (96.9%) | 1350 (97.1%) |
| Ethnicity | n=4,861 | n=3,410 | n=1,397 |
| White/Caucasian | 4454 (91.6%) | 3153 (92.5%) | 1264 (90.5%) |
| Black/African Canadian | 19 (0.4%) | 15 (0.4%) | ≤5 |
| Chinese | 19 (0.4%) | 14 (0.4%) | ≤5 |
| Indigenous | 11 (0.2%) | 7 (0.2%) | ≤5 |
| South Asian (Indian, Sri Lankan, etc.) | 17 (0.3%) | 7 (0.2%) | 9 (0.6%) |
| Southeast Asian (Japanese, Vietnamese, Korean, Cambodian, etc.) | 14 (0.3%) | 11 (0.3%) | ≤5 |
| West Asian (Arabian, Egyptian, Iranian, Afghan, etc.) | 10 (0.2%) | 7 (0.2%) | ≤5 |
| Other/Prefer to not say or self-identify | 317 (6.5%) | 196 (5.7%) | 106 (7.6%) |
| Language spoken most often at home | n=4,855 | n=3,411 | n=1,388 |
| English | 4627 (95.3%) | 3251 (95.3%) | 1327 (95.6%) |
| French | 165 (3.4%) | 120 (3.5%) | 41 (3.0%) |
| Other | 63 (1.3%) | 40 (1.2%) | 20 (1.4%) |
| Self-reported health status | n=4,873 | n=3,417 | n=1,397 |
| Excellent/very good/good | 4370 (89.7%) | 3082 (90.2%) | 1238 (88.6%) |
| Fair/poor | 492 (10.1%) | 330 (9.7%) | 154 (11.0%) |
| Don't Know | 11 (0.2%) | 5 (0.2%) | 5 (0.4%) |
| Location of residence<sup>b</sup> | n=4,752 | n=3,348 | n=1,354 |
| Urban | 3962 (83.4%) | 2791 (83.4%) | 1132 (83.6%) |
| Rural | 751 (15.8%) | 531 (15.9%) | 209 (15.4%) |
| Outside Canada | 39 (0.8%) | 26 (0.8%) | 13 (1.0%) |

<sup>a</sup> 61 respondents did not identify their gender
<sup>b</sup> 4405 (92.7%) respondents resided in Ontario and 308 (6.5%) in another Canadian province or territory.

Less than 5% (236/4790 [4.9%]) reported a cold or flu-like illness in the preceding month. Overall, 8 of 4861 respondents tested positive for COVID-19 (0.2%). Most respondents strongly agreed that the COVID-19 pandemic had changed their daily routine (67.5% females vs. 63.2% males, P=0.0047). Additional data on the impact of COVID-19 are reported in e**Table 1** and **eFigure 1** of the **Supplement**.

### Loneliness during COVID-19

Overall, 43.1% of respondents felt lonely at least some of the time, including 8.3% that felt lonely always or often (**Table 2**). Women were more likely to report feeling lonely than males (P<0.001). Strategies to avoid feeling lonely included connecting with a friend or family member (82.1% women vs. 70.7% men, P<0.001) and getting fresh air (65.3% vs. 61.9%, P=0.025). Seven percent (7.1%) described other strategies, such as reading, housework and/or gardening, and practising their faith. Most participants frequently spoke with a friend, family member or neighbour, although, a small proportion (0.4%) had no connection at all. Many used social networking websites (87.3% females vs. 78.2% males, P<0.001).

**Table 2.**
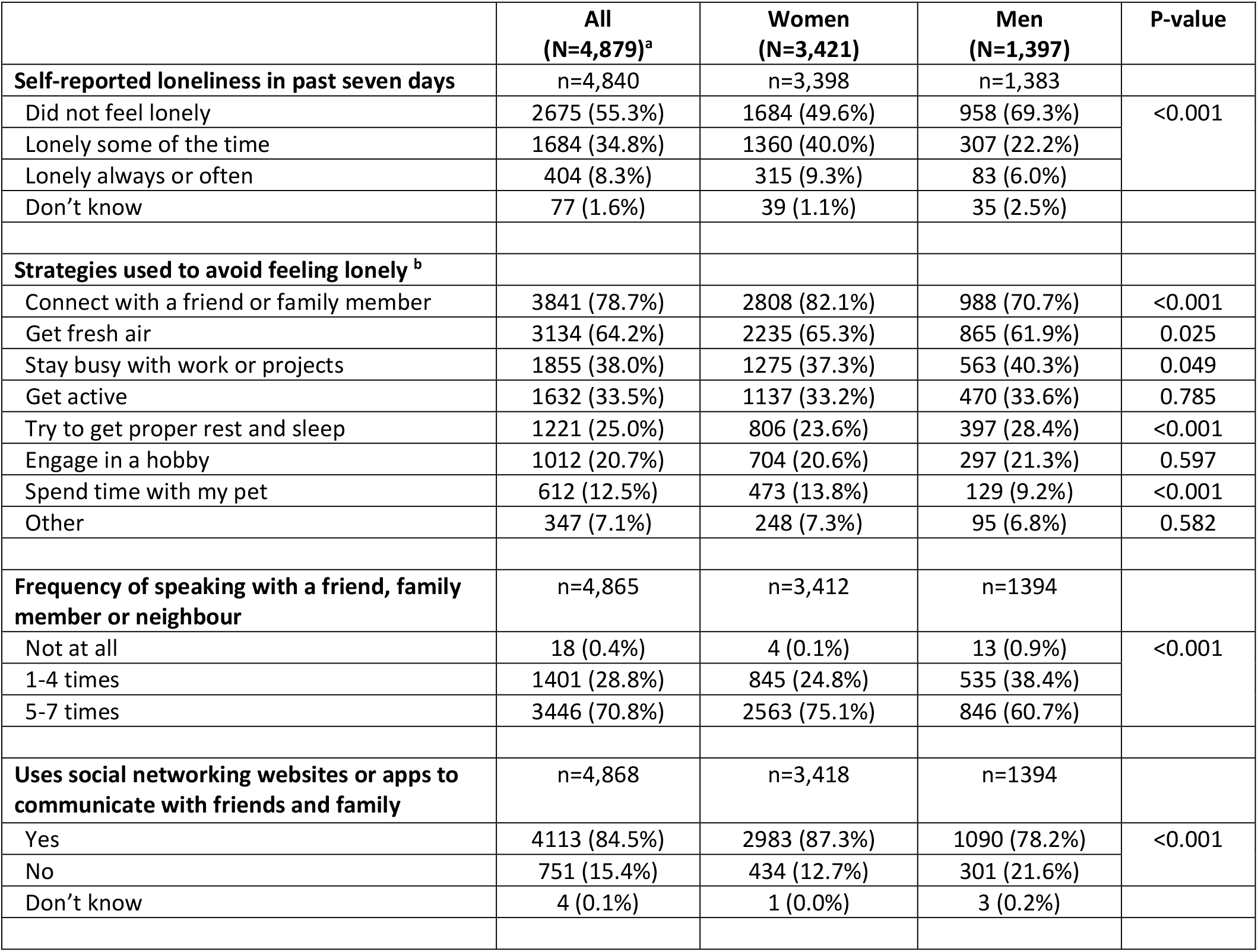
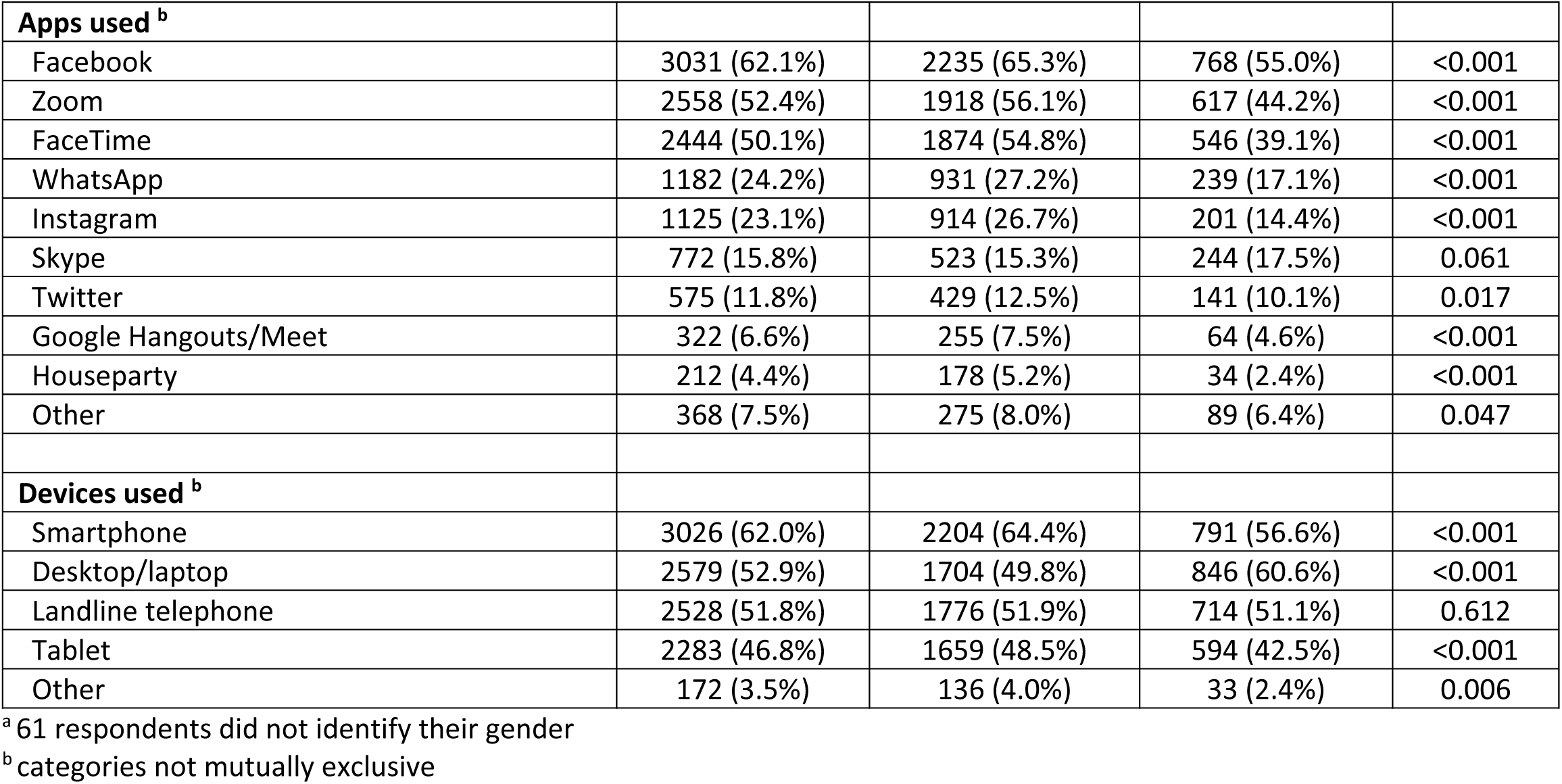
Loneliness and social connection in a sample of older Canadians, May 2020.

|  | **All**<br>**(N=4,879)<sup>a</sup>** | **Women**<br>**(N=3,421)** | **Men**<br>**(N=1,397)** | **P-value** |
| --- | --- | --- | --- | --- |
| **Self-reported loneliness in past seven days** | n=4,840 | n=3,398 | n=1,383 |  |
| Did not feel lonely | 2675 (55.3%) | 1684 (49.6%) | 958 (69.3%) | <0.001 |
| Lonely some of the time | 1684 (34.8%) | 1360 (40.0%) | 307 (22.2%) |  |
| Lonely always or often | 404 (8.3%) | 315 (9.3%) | 83 (6.0%) |  |
| Don't know | 77 (1.6%) | 39 (1.1%) | 35 (2.5%) |  |
| **Strategies used to avoid feeling lonely<sup>b</sup>** |  |  |  |  |
| Connect with a friend or family member | 3841 (78.7%) | 2808 (82.1%) | 988 (70.7%) | <0.001 |
| Get fresh air | 3134 (64.2%) | 2235 (65.3%) | 865 (61.9%) | 0.025 |
| Stay busy with work or projects | 1855 (38.0%) | 1275 (37.3%) | 563 (40.3%) | 0.049 |
| Get active | 1632 (33.5%) | 1137 (33.2%) | 470 (33.6%) | 0.785 |
| Try to get proper rest and sleep | 1221 (25.0%) | 806 (23.6%) | 397 (28.4%) | <0.001 |
| Engage in a hobby | 1012 (20.7%) | 704 (20.6%) | 297 (21.3%) | 0.597 |
| Spend time with my pet | 612 (12.5%) | 473 (13.8%) | 129 (9.2%) | <0.001 |
| Other | 347 (7.1%) | 248 (7.3%) | 95 (6.8%) | 0.582 |
| **Frequency of speaking with a friend, family member or neighbour** | n=4,865 | n=3,412 | n=1394 |  |
| Not at all | 18 (0.4%) | 4 (0.1%) | 13 (0.9%) | <0.001 |
| 1-4 times | 1401 (28.8%) | 845 (24.8%) | 535 (38.4%) |  |
| 5-7 times | 3446 (70.8%) | 2563 (75.1%) | 846 (60.7%) |  |
| **Uses social networking websites or apps to communicate with friends and family** | n=4,868 | n=3,418 | n=1394 |  |
| Yes | 4113 (84.5%) | 2983 (87.3%) | 1090 (78.2%) | <0.001 |
| No | 751 (15.4%) | 434 (12.7%) | 301 (21.6%) |  |
| Don't know | 4 (0.1%) | 1 (0.0%) | 3 (0.2%) |  |

**Table 2.**Loneliness and social connection in a sample of older Canadians, May 2020 (Continued)
|  | **All**<br>**(N=4,879)<sup>a</sup>** | **Women**<br>**(N=3,421)** | **Men**<br>**(N=1,397)** | **P-value** |
| --- | --- | --- | --- | --- |
| **Apps used <sup>b</sup>** |  |  |  |  |
| Facebook | 3031 (62.1%) | 2235 (65.3%) | 768 (55.0%) | <0.001 |
| Zoom | 2558 (52.4%) | 1918 (56.1%) | 617 (44.2%) | <0.001 |
| FaceTime | 2444 (50.1%) | 1874 (54.8%) | 546 (39.1%) | <0.001 |
| WhatsApp | 1182 (24.2%) | 931 (27.2%) | 239 (17.1%) | <0.001 |
| Instagram | 1125 (23.1%) | 914 (26.7%) | 201 (14.4%) | <0.001 |
| Skype | 772 (15.8%) | 523 (15.3%) | 244 (17.5%) | 0.061 |
| Twitter | 575 (11.8%) | 429 (12.5%) | 141 (10.1%) | 0.017 |
| Google Hangouts/Meet | 322 (6.6%) | 255 (7.5%) | 64 (4.6%) | <0.001 |
| Houseparty | 212 (4.4%) | 178 (5.2%) | 34 (2.4%) | <0.001 |
| Other | 368 (7.5%) | 275 (8.0%) | 89 (6.4%) | 0.047 |
| **Devices used <sup>b</sup>** |  |  |  |  |
| Smartphone | 3026 (62.0%) | 2204 (64.4%) | 791 (56.6%) | <0.001 |
| Desktop/laptop | 2579 (52.9%) | 1704 (49.8%) | 846 (60.6%) | <0.001 |
| Landline telephone | 2528 (51.8%) | 1776 (51.9%) | 714 (51.1%) | 0.612 |
| Tablet | 2283 (46.8%) | 1659 (48.5%) | 594 (42.5%) | <0.001 |
| Other | 172 (3.5%) | 136 (4.0%) | 33 (2.4%) | 0.006 |

<sup>a</sup> 61 respondents did not identify their gender
<sup>b</sup> categories not mutually exclusive

### Sex-stratified model

Most factors associated with loneliness were shared amongst women and men (**Table 3**). Older age significantly reduced the odds of loneliness in both sexes after adjustment for self-reported health status. Living alone was associated with loneliness in both women and men; although, the association was greater in men (adjusted Odds Ratio (aOR) 3.86 [95% Confidence Interval (CI) 2.88-5.18] vs. aOR 2.50 [95% CI 2.14-2.92]). Self-reported poor health and higher concern for the pandemic were also associated with loneliness, as were experiencing change to a daily routine, and not experiencing any positive effects or ‘silver linings’ of pandemic distancing measures; effect sizes varied by sex. Among women, receiving offers of assistance (aOR 0.79 [95% CI 0.69-0.91]) and communicating more often with a friend, family member or neighbour (aOR 0.47 [95% CI 0.34-0.66]) reduced the odds of loneliness.

**Table 3.** Odds ratios (OR) for loneliness stratified by sex in a sample of older Canadians, May 2020.

|  | Women |  |  | Men |  |  |
| --- | --- | --- | --- | --- | --- | --- |
|  | n (%)<br>Lonely | Unadjusted OR<br>(95% CI) | Age- & health-<br>adjusted OR<br>(95% CI) | n (%)<br>Lonely | Unadjusted OR<br>(95% CI) | Age- & health-<br>adjusted OR<br>(95% CI) |
| **Sociodemographic characteristics** |  |  |  |  |  |  |
| Age, years |  |  |  |  |  |  |
| <65 (ref) | 440 (52.8) | -- | -- | 65 (38.5) | -- | -- |
| 65-79 | 1110 (49.3) | 0.87 (0.74-1.02) | 0.84 (0.72-0.99) | 248 (27.1) | 0.59 (0.42-0.84) | 0.56 (0.39-0.78) |
| 80+ | 125 (46.3) | 0.77 (0.59-1.01) | 0.70 (0.53-0.92) | 77 (29.5) | 0.67 (0.45-1.01) | 0.61 (0.40-0.92) |
| Living arrangement |  |  |  |  |  |  |
| Lives with others (ref) | 935 (43.0) | -- | -- | 242 (23.0) | -- | -- |
| Lives alone | 714 (63.6) | 2.32 (2.00-2.67) | 2.50 (2.14-2.92) | 137 (54.2) | 3.95 (2.97-5.26) | 3.86 (2.88-5.18) |
| Ethnicity |  |  |  |  |  |  |
| White (ref) | 1565 (50.5) | -- | -- | 357 (29.2) | -- | -- |
| Non-White | 77 (41.6) | 0.70 (0.52-0.94) | 0.70(0.51-0.95) | 19 (26.4) | 0.87(0.51-1.49) | 0.83(0.48-1.43) |
| Residence of location |  |  |  |  |  |  |
| Urban (ref) | 1378 (50.4) | -- | -- | 312 (28.5) | -- | -- |
| Rural | 256 (48.7) | 0.94 (0.78-1.13) | 0.93 (0.77-1.13) | 58 (29.2) | 1.03 (0.74-1.44) | 1.09 (0.78-1.54) |
| Health status |  |  |  |  |  |  |
| Good (ref) | 1456 (48.1) | -- | -- | 324 (27.0) | -- | -- |
| Fair/Poor | 216 (66.9) | 2.18 (1.71-2.78) | 2.24 (1.76-2.86)* | 65 (45.1) | 2.22(1.56-3.16) | 2.34 (1.64-3.34)<sup>a</sup> |
| Caregiver to another person |  |  |  |  |  |  |
| No (ref) | 1198 (49.4) | -- | -- | 304 (28.5) | -- | -- |
| Yes | 469 (51.0) | 1.07 (0.92-1.25) | 1.05 (0.90-1.23) | 83 (30.1) | 1.08 (0.81-1.44) | 1.03 (0.77-1.39) |
| Receives care |  |  |  |  |  |  |
| No (ref) | 1447 (48.5) | -- | -- | 319 (27.5) | -- | -- |
| Yes | 220 (61.1) | 1.67 (1.33-2.09) | 1.55 (1.23-1.97) | 68 (37.6) | 1.59(1.15-2.20) | 1.39 (0.97-2.00) |

**Table 3.** Odds ratios (OR) for loneliness stratified by sex in a sample of older Canadians, May 2020 (Continued).
|  | Women |  |  | Men |  |  |
| --- | --- | --- | --- | --- | --- | --- |
|  | n (%)<br>Lonely | Unadjusted OR<br>(95% CI) | Age- & health-<br>adjusted OR<br>(95% CI) | n (%)<br>Lonely | Unadjusted OR<br>(95% CI) | Age- & health-<br>adjusted OR<br>(95% CI) |
| **Social Support** |  |  |  |  |  |  |
| Social media use |  |  |  |  |  |  |
| No (ref) | 213 (50.1) | -- | -- | 91 (31.5) | -- | -- |
| Yes | 1458 (49.8) | 0.99(0.80-1.21) | 1.00 (0.81-1.23) | 299 (28.4) | 0.86(0.65-1.14) | 0.90 (0.68-1.20) |
| Communication frequency<sup>b</sup> |  |  |  |  |  |  |
| None or low (ref) | 120 (68.6) | -- | -- | 55 (36.9) | -- | -- |
| High | 1551 (48.9) | 0.44 (0.32-0.61) | 0.47 (0.34-0.66) | 334 (27.9) | 0.66 (0.46-0.95) | 0.74 (0.61-1.06) |
| Received offers of assistance<sup>c</sup> |  |  |  |  |  |  |
| No (ref) | 1016 (52.5) | -- | -- | 253 (28.7) | -- | -- |
| Yes | 650 (46.3) | 0.78 (0.68-0.90) | 0.79 (0.69-0.91) | 136 (29.5) | 1.04 (0.81-1.33) | 1.05 (0.82-1.36) |
| **Attitudes and behaviours towards COVID-19** |  |  |  |  |  |  |
| Concern for pandemic |  |  |  |  |  |  |
| Low level (ref) | 260 (42.1) | -- | -- | 62 (19.8) | -- | -- |
| High level | 1407 (51.6) | 1.47 (1.23-1.75) | 1.46 (1.22-1.74) | 328 (31.8) | 1.90 (1.40-2.58) | 1.86 (1.36-2.53) |
| Extent practising physical distancing |  |  |  |  |  |  |
| None/some (ref) | 155 (47.3) | -- | -- | 40 (22.5) | -- | -- |
| Most of time | 1231 (49.9) | 1.11(0.88-1.40) | 1.06 (0.84-1.34) | 295 (29.9) | 1.47(1.01-2.15) | 1.41 (0.96-2.07) |
| All of time | 283 (51.4) | 1.18 (0.90-1.55) | 1.06(0.80-1.40) | 55 (30.7) | 1.53 (0.95-2.46) | 1.31 (0.80-2.14) |
| No perceived positive effects of distancing |  |  |  |  |  |  |
| No (ref) | 1331 (46.7) | -- | -- | 306 (27.5) | -- | -- |
| Yes | 344 (67.3) | 2.35(1.92-2.86) | 2.25 (1.84-2.75) | 84 (35.9) | 1.48(1.10-1.99) | 1.44 (1.06-1.95) |

**Table 3.** Odds ratios (OR) for loneliness stratified by sex in a sample of older Canadians, May 2020 (Continued).
|  | Women |  |  | Men |  |  |
| --- | --- | --- | --- | --- | --- | --- |
|  | n (%)<br>Lonely | Unadjusted OR<br>(95% CI) | Age- & health-<br>adjusted OR<br>(95% CI) | n (%)<br>Lonely | Unadjusted OR<br>(95% CI) | Age- & health-<br>adjusted OR<br>(95% CI) |
| Change in daily routine |  |  |  |  |  |  |
| No (ref) | 46 (34.9) | -- | -- | 6 (8.2) | -- | -- |
| Yes | 1623 (50.4) | 1.90 (1.32-2.74) | 2.02 (1.39-2.92) | 383 (30.2) | 4.83(2.08-11.24) | 5.57(2.37-13.11) |

<sup>a</sup> Adjusted for age group only.
<sup>b</sup> Self-reported communication with friends, family members or neighbours.
<sup>c</sup> Reported receiving offers of assistance from their community to help with daily life during COVID-19 distancing measures.

### Sex-pooled model

Women had increased odds of loneliness compared to men, irrespective of living arrangement (aOR 1.52 [95% CI 1.13-2.04] living alone; aOR 2.44 [95%CI 2.04-2.92] living with others) (**Table 4**). Increasing age group was associated with decreasing odds of loneliness. The association of living alone with loneliness was significantly greater for men than women (aOR 4.26 [95% CI 3.15-5.76] vs. 2.65 [95% CI 2.26-3.11], P=0.006 for interaction term). Additional characteristics associated with loneliness included: self-reported fair/poor health (aOR 1.93 [95% CI 1.54-2.41]), being a caregiver (aOR 1.18 [95% CI 1.02-1.37]) and receiving care from a caregiver (aOR 1.47 [95% CI 1.19-1.81]). Pandemic-related factors associated with an increased odds of loneliness included having a high concern for the pandemic (Aor 1.55 [95% CI 1.31-1.84]), not experiencing any positive effects or ‘silver linings’ of pandemic distancing measures (aOR 1.94 [95% CI 1.62-2.32]) and experiencing change to a daily routine (aOR 2.81 [95% CI 1.96-4.03]). Non-white ethnicity (aOR 0.71 [95% CI 0.54-0.94]), high frequency of communication (aOR 0.55 [95% CI 0.43-0.72]) and receiving offers of assistance (aOR 0.79 [95% CI 0.69-0.90]) reduced the odds of loneliness. None of the other sex-based interactions we explored with explanatory factors were significant.

**Table 4.** Odds ratios (OR) for loneliness (sex-pooled) in a sample of older Canadians, May 2020.

|  | All respondents |  |  |  |
| --- | --- | --- | --- | --- |
|  | Unadjusted OR (95% CI) | Age- & sex- Adjusted OR (95% CI) | Age-, sex-, & health status- Adjusted OR (95% CI) | Fully<sup>a</sup> adjusted OR (95% CI) |
| **Sociodemographic** |  |  |  |  |
| Female sex (ref male) | 2.44 (2.13-2.80) | 2.38 (2.07-2.73) | 2.41 (2.09-2.77) |  |
| Women living alone |  |  |  | 1.52 (1.13-2.04) |
| Women living with others |  |  |  | 2.44 (2.04-2.92) |
| Age, years |  |  |  |  |
| 65-79 (ref <65) | 0.74 (0.64-0.86) | 0.81 (0.70-0.94) | 0.78 (0.67-0.90) | 0.69 (0.59-0.81) |
| 80+ (ref <65) | 0.61 (0.49-0.75) | 0.79 (0.63-0.98) | 0.72 (0.57-0.90) | 0.50 (0.39-0.65) |
| Living alone | 2.83 (2.49-3.22) | 2.78 (2.42-3.18) | 2.74 (2.39-3.15) |  |
| Living alone in women |  |  |  | 2.65 (2.26-3.11) |
| Living alone in men |  |  |  | 4.26 (3.15-5.76) |
| Non-white ethnicity | 0.75 (0.58-0.97) | 0.74 (0.57-0.96) | 0.72 (0.55-0.94) | 0.71 (0.54-0.94) |
| Rural | 0.98 (0.83-1.15) | 0.95 (0.81-1.12) | 0.96 (0.82-1.13) | 1.07 (0.90-1.27) |
| Fair or poor health status | 2.14 (1.76-2.60) | 2.25 (1.84-2.76) | -- | 1.93 (1.54-2.41) |
| Caregiver to another person | 1.14 (1.00-1.30) | 1.04 (0.91-1.20) | 1.05 (0.91-1.20) | 1.18 (1.02-1.37) |
| Receives care | 1.54 (1.29-1.84) | 1.76 (1.45-2.12) | 1.50 (1.24-1.83) | 1.47 (1.19-1.81) |
| **Social support** |  |  |  |  |
| Social media use | 1.08 (0.92-1.26) | 0.93 (0.78-1.09) | 0.96 (0.81-1.14) | 1.13 (0.94-1.36) |
| High communication frequency | 0.65 (0.52-0.81) | 0.53 (0.42-0.68) | 0.57 (0.45-0.72) | 0.55 (0.43-0.72) |
| Received offers of assistance | 0.89 (0.79-1.00) | 0.85 (0.75-0.96) | 0.85 (0.75-0.96) | 0.79 (0.69-0.90) |
| **Attitudes and behaviours towards COVID-19** |  |  |  |  |
| High concern for pandemic | 1.65 (1.42-1.91) | 1.59 (1.37-1.86) | 1.56 (1.33-1.82) | 1.55 (1.31-1.84) |
| Extent practising distancing |  |  |  |  |
| Most of time (ref none/some) | 1.27 (1.05-1.53) | 1.19 (0.98-1.45) | 1.15 (0.95-1.40) | 1.23 (0.99-1.53) |
| All of time (ref none/some) | 1.39 (1.11-1.75) | 1.29 (1.02-1.64) | 1.13 (0.89-1.44) | 1.12 (0.86-1.45) |
| No perceived positive effects of pandemic distancing measures | 1.90 (1.62-2.22) | 2.07 (1.76-2.43) | 1.97 (1.67-2.32) | 1.94 (1.62-2.32) |
| Reported change in routine | 2.36 (1.72-3.24) | 2.30 (1.67-3.19) | 2.50 (1.80-3.48) | 2.81 (1.96-4.03) |

<sup>a</sup> Adjusted for all covariates listed in the table with an interaction term for sex and living alone (P-value =0.006).

## Discussion

In a survey of 4879 older women and men, we found that loneliness was common during the COVID-19 pandemic, with more than one-third (34.8%) of respondents reporting feeling lonely sometimes and 8.3% feeling lonely always or often. More women reported feeling lonely than men and had higher odds of loneliness, despite controlling for factors hypothesized to contribute to sex differences including living alone, health status, and caregiving. Our findings are similar to reports from the UK, where 22.4% and 4.1% of older adults reported feeling lonely sometimes or often, respectively, in the first four weeks of lockdown^27^, and from the US, where 13.8% (95% CI 11.4%-16.6%) of adults aged ≥18 years reported feeling lonely always or often at the beginning of April 2020.^22^

Living alone is as an important risk factor for loneliness, both pre-COVID-19^4,28,29^ and presently.^27,30,31^ We found that living alone predicted loneliness in women and men, although the effect was greater in men. Physical distancing and stay-at-home measures are anticipated to have a greater toll for those living alone as they severely limit opportunities for face-to-face interaction to combat loneliness.^30^ The effect of living alone on loneliness may be greater in men because they tend to have fewer social contacts and close friends than women.^32-34^ Indeed, male respondents in our survey communicated less frequently with family, friends, and neighbours, and were less likely to seek out social connection to mitigate loneliness. Having a smaller social network may exacerbate some of the negative effects of living alone. Emerson recently found that older US adults who lived alone were less likely to have a close relationship that provided emotional security and well-being, and more likely to become ‘more lonely’ following the onset of COVID-19than those living with others (42.4% vs. 27.9%).^31^ Alternatively, our finding may be due to the inherent overlap in the constructs of ‘living alone’ and ‘marital status’ because we partially captured the impact of being widowed or unmarried in men versus women. Prior research has shown that being single has a greater impact on men’s loneliness, possibly explained by the fact that for many older men, their partners are their main confidante and source of intimacy.^35,36^

We found that older adults’ perceptions and pandemic experiences were also associated with loneliness. Respondents who had a high level of concern for COVID-19, experienced changes to their daily routine, and reported no perceived positive effects or ‘silver livings’ from the pandemic had increased odds of loneliness, while receiving offers of support and frequently communicating with family, friends and neighbours were protective. These findings underscore the importance of public health messages from the World Health Organization targeted at older adults, including maintaining regular routines or creating new ones that include exercise, regular cleaning/chores, and enjoyable activities; keeping in regular contact with loved ones; and restricting news consumption to specific times of day from reputable sources to reduce undue anxiety or distress.^37^

Family physician visits have been suggested as an important opportunity to screen for loneliness during COVID-19.^38,39^ Particular attention is recommended to be paid to patients who are older, live alone or have pre-existing health conditions.^38^ Our findings suggest that considering the patient’s sex, if they have sufficient social support, and how the pandemic is affecting their daily routines could further assist in identifying at-risk individuals. Such questions would also be beneficial to align patients more purposefully with interventions. Virtual consultations and social prescribing (i.e. linking patients with nonclinical supports in their community such as outdoor exercise classes, walking groups, virtual bereavement programs, etc) may be effective strategies to reduce loneliness during COVID-19 and beyond. ^38,40,41^

Lastly, digital technologies and platforms can facilitate social connection;^40,42^ although, recent research shows that many older adults lack access to internet-enabled devices^43^, and are unready for comparable technologies (i.e. video telemedicine visits) due to inexperience with technology or physical disability.^44^ Consistent with prior research^31,45^ and likely a function of electronic survey administration, we found high levels (∼85%) of social media engagement, with no increased risk for loneliness. Our findings suggest there is a large segment of the older adult population for whom digital media-based interventions may be effective for mitigating and alleviating loneliness. Services that teach older adults how to use and connect with family and friends through social media platforms may be valuable.^46^ The importance of offline connection, however, should not be forgotten – phoning parents or older neighbours, and extending offers of assistance can go a long way to making someone feel connected and visible.^47^

A recent US study reported that 30.9% of older adults surveyed felt more lonely after COVID-19 related physical distancing was implemented.^31^ Our estimates of loneliness were almost double that of the CLSA’s collected between 2010-2015 using a similar age group and measurement approach (49.3% of women and 27.1% men aged 65-79 years felt lonely some of the time vs. 24.7% and 17.9%, respectively, for adults aged 65-74 years).^2,48^ Comparisons should, be made cautiously considering differences in study populations. Longitudinal studies provide the most robust evidence of temporal changes. Using data collected at three time points, Luchetti et al found that older adults were the only group studied that showed a slight increase in loneliness in late March 2020 after social distancing measures were implemented in the US compared to the baseline assessment in January/February, although levels remained stable in April.^30^ The study found that this increase was driven primarily by unavailable social connections, rather than feelings of isolation. It will continue to be important to consistently measure how rates of loneliness change over the course of the pandemic to identify drivers and determine at-risk populations who could benefit from additional support.

### Limitations

Our study leveraged a strong community-based partnership to obtain timely data from a large sample of older Canadians on the impacts of COVID-19. Analyses were exploratory and intended to identify characteristics and circumstances associated with loneliness to help target supports to those who could benefit from them. A limitation is that the data are based on a convenience sample of retired, educational staff, who are not fully representative of the Canadian population. The perspectives of vulnerable groups who may be at greater risk for loneliness (e.g. those with severe mental health illness, low income, no home internet access, etc.) are likely underrepresented in this sample. As such, our findings may be a conservative estimate of loneliness.

### Conclusions

While many older adults reported feeling lonely during COVID-19, several characteristics – in particular being female and living alone – increased the odds of loneliness. These characteristics may help guide targeting interventions to reduce loneliness as the pandemic persists.

## Data Availability

The dataset used for this analysis is not publicly available. The analytic code is available from the authors upon request

## Acknowledgements

Study authors thank RTOERO staff who assisted in the survey and members who completed the survey.

## Funding

Dr Savage is supported by a Canadian Institutes of Health Research Postdoctoral Fellowship [MFE 158218]. Dr Chamberlain is supported by a Canadian Institutes of Health Postdoctoral Fellowship. Dr Stall receives funding from the Canadian Institutes of Health Research Vanier Scholarship Program, the Eliot Phillipson Clinician-Scientist Training Program and the Clinician Investigator Program at the University of Toronto. Dr Rochon is the RTOERO Chair in Geriatric Medicine at the University of Toronto.

## Author Contributions

*Study concept and design*: Savage, Rochon

*Acquisition, analysis, or interpretation of data*: All authors.

*Drafting of the manuscript*: Savage

*Statistical analysis*: Wu

*Critical revision of the manuscript for important intellectual content*: All authors.

## Role of the Funder/Sponsor

Study funders/sponsors had no role in the design and conduct of the study; collection, management, analysis, and interpretation of the data; preparation, review, or approval of the manuscript; nor the decision to submit the manuscript for publication.

## Competing interests

None declared

## Notes

### Competing Interest Statement

The authors have declared no competing interest.

### Funding Statement

No specific funding was received. Dr Savage is supported by a Canadian Institutes of Health Research Postdoctoral Fellowship [MFE 158218]. Dr Chamberlain is supported by a Canadian Institutes of Health Postdoctoral Fellowship. Dr Stall receives funding from the Canadian Institutes of Health Research Vanier Scholarship Program, the Eliot Phillipson Clinician-Scientist Training Program and the Clinician Investigator Program at the University of Toronto. Dr Rochon is the RTOERO Chair in Geriatric Medicine at the University of Toronto.

### Author Declarations

The Research Ethics Board at Women's College Hospital in Toronto, Canada approved this study [#2020-0051-E].

