## Supplemental Files for "Loneliness among older adults in the community during COVID-19"

eAppendix. Questionnaire

eMethods. Exposure Variable Definitions

eTable 1. Impact of COVID-19 on daily life reported by a sample of older Canadians, May 2020

eFigure 1. Top new or additional concerns related to COVID-19 and physical distancing measures reported by survey respondents, May 2020.

### eAppendix. Questionnaire

The Impact of COVID-19 Physical Distancing Measures on Older Canadians and Strategies to Address Unmet Needs: A Survey of Retired Educators

#### Introduction

Welcome! Thank you for agreeing to participate in this survey. We value your opinions and we appreciate your participation in this process.

The Study Information Sheet will answer many of your questions and reviews your rights and responsibilities as a participant in this research project. You can access the Study Information Sheet by clicking this link. You may print a copy of the Study Information Sheet for your records.

If you have additional questions, please contact Joyce Li, Research Coordinator before continuing further.

#### Electronic Consent

Please select your choice below. Clicking on the "Agree" button indicates your confirmation that:

This research study has been fully explained to me and all of my questions answered to my satisfaction
I understand the requirements of participating in this research study
I have been informed of the risks and benefits, if any, of participating in this research study
I have been informed of any alternatives to participating in this research study
I have been informed of the rights of research participants
I have read each page of the Study Information Sheet
I have agreed to participate in this research study

**Electronic Consent**

- ○ Agree
- ○ Disagree

The Coronavirus pandemic (COVID-19) is impacting all Canadians but older adults are experiencing its impacts in unique ways. This survey will help us understand if and how COVID-19 is affecting your health, as well your social circumstances and supports you have available. This information will be used by researchers at Women's College Hospital as well as RTOERO leadership to develop supports for older adults and for our members during and after the COVID-19 pandemic. The survey is anonymous and will take about 10-20 minutes to complete.

#### A) Daily life during COVID-19

**1. To what extent would you agree with the following statement: The Covid-19 crisis has changed my daily routine.**

- ○ Strongly Agree
- ○ Somewhat Agree
- ○ Neutral
- ○ Somewhat Disagree
- ○ Strongly Disagree

○ Don’t know

Comment:

2. How are you spending your time now? Select all that apply.

◻ Watching more TV
◻ More time on my hobbies
◻ COVID-19-related community work (making masks, grocery shopping, meal or supply drop-offs, etc)
◻ Working from home
◻ Going on walks
◻ More time exercising
◻ More time cooking or baking
◻ More time making or taking phone calls from friends/relatives
◻ More time on the internet and social media
◻ I am not spending my time differently than before COVID-19
◻ Other, please specify:

3. Have you experienced any of the following difficulties due to COVID-19? Please select all that apply.

◻ Getting/ordering groceries
◻ Getting supplies (e.g. toilet paper, hand sanitizer, cleaning products, bleach, etc.)
◻ Getting prescription medications
◻ Accessing healthcare
◻ Changes to planned health treatments (e.g. cancer treatment, outpatient procedure, surgery, etc). Please Specify:
◻ Other, please describe:
◻ I have not experienced any difficulties

4. Although this is a challenging time, have you experienced any positive effects or 'silver linings' during this crisis? Please select all that apply.

◻ Stronger sense of community
◻ Feeling more connected to partner, family and friends
◻ A growing respect for older adults and their needs by society (e.g. designated grocery shopping hours)
◻ Slower pace of life / more time to relax or rest
◻ No or less time spent commuting to work
◻ Improved access to healthcare through virtual care
◻ Other, please describe:
◻ I have not experienced any positive effects of this crisis

Comment:

5. How concerned are you about the COVID-19 pandemic?

○ Extremely concerned
○ Very concerned
○ Moderately concerned
○ Slightly concerned
○ Not at all concerned

6. To what extent are you practising physical distancing?

- ○ All of the time. I am staying home all of the time.
- ○ Most of the time. I only leave my home to buy essentials or for necessary medical appointments.
- ○ Some of the time. I have reduced the amount of time I spend in public.
- ○ None of the time. I am doing everything that I normally do.

7. The COVID-19 pandemic and physical distancing measures have created new or additional concerns for many people. Select your top three concerns.

- □ Getting sick from COVID-19
- □ A loved one getting sick from COVID-19
- □ The health system becoming overloaded (not enough hospital beds or supplies)
- □ Not being able to meet basic needs (put food on the table or pay bills)
- □ Feeling lonely, anxious or depressed
- □ Limited access to routine healthcare
- □ Not being able to adequately take care of my health
- □ Not being able to adequately care for loved ones
- □ Not being able to visit loved ones in long-term care
- □ Family stress from confinement
- □ Unwittingly spreading COVID-19 (if sick without symptoms)
- □ My children or grandchildren's education or work
- □ Economic recession and retirement savings
- □ Other – please indicate:

8. In the past 4 weeks, have you been in close contact with a person who has tested positive for COVID-19?

- ○ Yes
- ○ No
- ○ Don't know

9. In the past 4 weeks, have you been ill with a cold or flu-like illness?

- ○ Yes
- ○ No
- ○ Don't know

10. Have you been tested for COVID-19?

- ○ Yes, I was tested and was positive
- ○ Yes, I was tested and was negative
- ○ No, I tried to get tested but could not get a test
- ○ No, I have not tried to get tested

#### B) Caregiving and receiving care

11. Do you provide assistance to another person because of a health condition or limitation? By assistance we mean personal care, medical treatments, scheduling or coordinating care-related tasks, meal preparation, house maintenance, transportation, social or emotional support, mobility, or financial assistance or management. Please exclude any assistance you provided as part of a volunteer organization or paid job.

- ○ Yes
- ○ No

○ Don’t Know

Do you live in the same household as this person?

○ Yes
○ No
○ Don’t Know

Has the COVID-19 crisis impacted your ability to give care? In what way?

○ Yes, please specify:
○ No
○ Don’t know

Comment:

12. Do you receive assistance from family, friends, or neighbours because of a health condition or limitation that affects your daily activities?

○ Yes
○ No
○ Don’t Know

Does your caregiver live in the same household as you?

○ Yes
○ No
○ Don’t Know

Has the COVID-19 crisis impacted your ability to receive care? In what way?

○ Yes, please specify:
○ No
○ Don’t know

#### C) Social connections during COVID-19

To reduce the spread of COVID-19, the government and public health officials have asked Canadians to practise physical distancing (i.e. minimizing close contact with others). While physical distancing is necessary to slow the spread of disease, it may lead to loneliness, anxiety or depression.

13. In the past seven days, which statement best applies?

○ I did not feel lonely.
○ I felt lonely one or two days.
○ I felt lonely several days.
○ I felt lonely most days.
○ I felt lonely every day.
○ Don’t know.

Comment:

14. What steps do you take to avoid feeling lonely? Please select up to three strategies you use most often.

- ◻ Connect with a friend or family member
- ◻ Get fresh air
- ◻ Get active
- ◻ Stay busy with work or projects
- ◻ Engage in a hobby
- ◻ Try to get proper rest and sleep
- ◻ Spend time with my pet
- ◻ Other, please share any strategies:
- ◻ Please share with us any specific resources you use to avoid feeling lonely (e.g., participating in a virtual book club):

15. In the past seven days, how often did you speak with a friend, family member or neighbour?

- ○ Not at all
- ○ 1-2 times
- ○ Several times (3-4 times)
- ○ Almost every day (5-6 times)
- ○ Every day (7 times)

#### D) Use of technology to stay socially connected

Digital technologies can help us stay socially connected as we practise physical distancing.

16. Do you have access to the Internet at home?

- ○ Yes
- ○ No
- ○ Don't Know

What are the reasons you do not have access to the internet at home? Select all that apply.

- ◻ No need or no interest
- ◻ Cost (service or equipment)
- ◻ The available service does not meet our needs
- ◻ Security or privacy concerns (e.g. viruses, use of personal information)
- ◻ Lack of confidence, knowledge, or skills
- ◻ No Internet-ready device (e.g. desktop computer) available in household
- ◻ Other, please specify:

How would you rate the internet connection in your home?

- ○ Very good
- ○ Good
- ○ Moderate
- ○ Poor
- ○ Don't know

17. Do you have a smartphone that you use for personal use? A mobile phone that performs many of the functions of a computer, typically having a touchscreen interface, Internet access, and an operating system capable of running downloaded applications, e.g. Apple iPhone and Samsung Galaxy

- ○ Yes
- ○ No

○ Don’t know

18. Do you use any social networking websites (e.g. Facebook) or apps (e.g. Zoom or FaceTime) to communicate with friends and family?

○ Yes
○ No
○ Don’t know

Please check which sites or apps you use (check all that apply)

□ Facebook
□ Instagram
□ Twitter
□ WhatsApp Messenger
□ Zoom
□ Skype
□ Face Time
□ Houseparty
□ Google Hangouts/meet
□ Other, please specify:

19. What devices do you use most often when connecting with friends and family? Please select all that apply.

□ Desktop/Laptop
□ Tablet
□ Smartphone
□ Landline telephone
□ Other, please specify:

Comment:

#### E) Supporting older adults during the COVID-19

20. In your view, what are the most pressing needs of older adults during the COVID-19 pandemic? Please select up to 3 issues.

□ Support for caregivers
□ Access to (routine?) healthcare to maintain physical health
□ Resources or supports on how to stay physically healthy during the COVID-19
□ Resources or supports on how to stay mentally healthy during the COVID-19
□ Programs or supports to ensure basic needs are met (e.g. foodbanks, home meal delivery, income supplements, etc.)
□ Policies and procedures to ensure safety of older adults in long-term care
□ Strategies to ensure older adults are able to stay connected with loved ones in long-term care
□ Strategies to help older adults stay socially connected while physically distanced
□ Other, please specify:

Comment:

21. To what extent do you agree or disagree with the following statements?

| | Strongly agree | Somewhat agree | Neutral | Somewhat disagree | Strongly disagree |
|---|---|---|---|---|---|
| **a. I have received offers of assistance from my community to help with daily life during stay at home and physical distancing measures.** | ○ | ○ | ○ | ○ | ○ |
| **b. Governments and policy makers care about the health and well-being of older adults.** | ○ | ○ | ○ | ○ | ○ |
| **c. The level of respect for older adults in society has decreased during the COVID-19 pandemic.** | ○ | ○ | ○ | ○ | ○ |
| **d. I have witnessed ageism in the daily news and popular culture during the COVID-19 pandemic.** | ○ | ○ | ○ | ○ | ○ |

Comment:

#### F) Sociodemographics

23. Your age

- ○ 54 or younger
- ○ 55-59
- ○ 60-64
- ○ 65-69
- ○ 70-74
- ○ 75-79
- ○ 80+

24. Your gender

- ○ Female
- ○ Male
- ○ Prefer to self identify
- ○ Prefer not to say

25. Including yourself, how many persons are living in your household?

26. Do you have access to private outdoor space (e.g. backyard, terrace or balcony)?

- ○ Yes
- ○ No
- ○ Don’t Know

27. How would you describe your ethnic identity?

- ○ Black/African Canadian
- ○ Central/South American

- Chinese
- Filipino
- Indigenous
- South Asian (Indian, Sri Lankan, etc.)
- Southeast Asian (Japanese, Vietnamese, Korean, Cambodian, etc.)
- West Asian (Arabian, Egyptian, Iranian, Afghan, etc.)
- White/Caucasian (European, Russian, etc.)
- Other, please specify:
- Prefer to self-identify
- Prefer not to say

28. What language do you speak most often at home?

- English
- French
- Other, please indicate:

29. In general, would you say your health is... ?

- Excellent
- Very good
- Good
- Fair
- Poor
- Don’t Know

30. What are the first 3 digits of your postal code?

#### G) Overall comments and suggestions

31. How can RTOERO and the Foundation support members during the COVID-19 pandemic?

32. Other comments or suggestions

You have opted not to consent to participate at this time. Thank you for considering the invitation to participate in this survey project.

#### eMethods. Exposure Variable Definitions

| Sociodemographic | Definition |
|---|---|
| Sex | Based on self-identification as female or male. |
| Age | Categorized as <65 years if respondent's selected age was '54 or younger', '55-59', or '60-64'; as 65-79 years if they selected '65-69', '70-74' or '75-79'; and as 80+ if they selected '80+'. |
| Living arrangement | Classified as living alone if reported 1 person living in their household (i.e. themselves) and as living with others if reported >1 person living in their household. |
| Ethnicity | Classified as white if respondents identified themselves as 'White/Caucasian' or they identified as 'Other' but specified white, Caucasian, Hebrew/Jewish, or white European ethnicity, e.g. Italian, French, Irish, Greek, Welsh, Scottish, etc. Central/South American and Filipino were regrouped into the Other category due to small numbers. |
| Rural residence | Classified as rural if second digit of reported Canadian postal code was a '0', and outside Canada if no match to a Canadian postal code.[1] |
| Health status | Classified as 'fair or poor' based on self-reporting fair or poor health; and as 'good' if 'excellent', 'very good' or 'good' health was reported. |
| Caregiver | Classified as a caregiver if responded that they aid another person because of a health condition or limitation. |
| Care recipient | Classified as a care recipient if they reported receiving assistance from another person because of a health condition or limitation. |
| **Social support** | |
| Social media use | Classified as yes if respondent reported using any social networking websites (e.g. Facebook) or apps (e.g. Zoom or FaceTime) to communicate with friends and family. |
| Frequency of communication | Classified as 'high frequency' if reported speaking with a friend, family member or neighbour≥3 times in the prior week. |
| Receipt of offers of assistance | Classified as yes if respondent strongly or somewhat agreed to the statement "I have received offers of assistance from my community to help with daily life during stay at home and physical distancing measures." |
| **Attitudes and behaviours towards COVID-19** | |
| Level of concern | Classified as 'high concern' if respondent reported they were 'extremely' or 'very concerned' about the COVID-19 pandemic. |
| Extent practicing physical distancing | Classified as 'all of the time', 'most of the time' or 'some of the time or none' based on self-report. |
| Change in routine | Classified as yes if respondent strongly or somewhat agreed that the Covid-19 crisis changed their daily routine, and as no if respondent was neutral, or somewhat or strongly disagreed with the statement. |

#### eTable 1. Impact of COVID-19 on daily life reported by a sample of older Canadians, May 2020

| | All (N=4,879)[a] | Female (N=3,421) | Male (N=1,397) | P-Value |
|---|---|---|---|---|
| **The COVID-19 crisis has changed my daily routine** | n=4,863 | n=3,412 | n=1,390 | |
| Strongly Agree | 3211 (66.0%) | 2304 (67.5%) | 878 (63.2%) | 0.0047 |
| Somewhat Agree | 1438 (29.6%) | 973 (28.5%) | 436 (31.4%) | |
| Neutral | 91 (1.9%) | 56 (1.6%) | 35 (2.5%) | |
| Somewhat Disagree | 87 (1.8%) | 60 (1.8%) | 25 (1.8%) | |
| Strongly Disagree | 35 (0.7%) | 18 (0.5%) | 16 (1.2%) | |
| Don't know | 1 (0.0%) | 1 (0.0%) | 0 | |
| **How time is being spent [b]** | | | | |
| More time on the internet and social media | 3584 (73.5%) | 2562 (74.9%) | 978 (70.0%) | 0.0005 |
| Going on walks | 3128 (64.1%) | 2260 (66.1%) | 835 (59.8%) | <0.0001 |
| Watching more TV | 2877 (59.0%) | 2039 (59.6%) | 805 (57.6%) | 0.2050 |
| More time making or taking phone calls from friends/relatives | 2593 (53.2%) | 2026 (59.2%) | 543 (38.9%) | <0.0001 |
| More time cooking or baking | 2517 (51.6%) | 2001 (58.5%) | 489 (35.0%) | <0.0001 |
| More time on my hobbies | 2073 (42.5%) | 1527 (44.6%) | 518 (37.1%) | <0.0001 |
| More time exercising | 1111 (22.8%) | 780 (22.8%) | 320 (22.9%) | 0.9367 |
| COVID-19-related community work | 592 (12.1%) | 500 (14.6%) | 83 (5.9%) | <0.0001 |
| Working from home | 431 (8.8%) | 291 (8.5%) | 136 (9.7%) | 0.1733 |
| Other | 987 (20.2%) | 691 (20.2%) | 283 (20.3%) | 0.9631 |
| Cleaning, home renovations, gardening, organizing/decluttering | 308 (6.3%) | | | |
| Reading | 198 (4.1%) | | | |
| Not spending my time differently than before COVID-19 | 179 (3.7%) | 89 (2.6%) | 86 (6.2%) | <0.0001 |
| **Difficulties experienced [b]** | | | | |
| Getting supplies (e.g. toilet paper, hand sanitizer, cleaning products, bleach, etc.) | 2029 (41.6%) | 1471 (43.0%) | 528 (37.8%) | 0.0009 |
| Getting/ordering groceries | 1611 (33.0%) | 1130 (33.0%) | 459 (32.9%) | 0.9066 |
| Changes to planned health treatments (e.g. cancer treatment, outpatient procedure, surgery, etc). | 1296 (26.6%) | 890 (26.0%) | 388 (27.8%) | 0.2098 |
| Accessing healthcare | 1040 (21.3%) | 697 (20.4%) | 326 (23.3%) | 0.0226 |
| Getting prescription medications | 687 (14.1%) | 448 (13.1%) | 230 (16.5%) | 0.0023 |
| Other | 776 (15.9%) | 602 (17.6%) | 171 (12.2%) | <0.0001 |
| Prescription, medications on backorder | 40 (0.8%) | | | |
| No difficulties experienced | 1353 (27.7%) | 939 (27.5%) | 398 (28.5%) | 0.4638 |

#### eTable 1. Impact of COVID-19 on daily life reported by a sample of older Canadians, May 2020 (Continued)

| | All (N=4,879)[a] | Female (N=3,421) | Male (N=1,397) | P-Value |
|---|---|---|---|---|
| **Positive effects experienced [b]** | | | | |
| Slower pace of life / more time to relax or rest | 2583 (52.9%) | 1879 (54.9%) | 673 (48.2%) | <0.0001 |
| Feeling more connected to partner, family and friends | 2062 (42.3%) | 1405 (41.1%) | 629 (45.0%) | 0.0117 |
| A growing respect for older adults and their needs by society | 1778 (36.4%) | 1279 (37.4%) | 473 (33.9%) | 0.0209 |
| Stronger sense of community | 1571 (32.2%) | 1129 (33.0%) | 429 (30.7%) | 0.1225 |
| No or less time spent commuting to work | 341 (7.0%) | 240 (7.0%) | 96 (6.9%) | 0.8590 |
| Improved access to healthcare through virtual care | 190 (3.9%) | 143 (4.2%) | 47 (3.4%) | 0.1868 |
| Other | 492 (10.1%) | 374 (10.9%) | 113 (8.1%) | 0.0030 |
| None experienced | 778 (16.0%) | 519 (15.2%) | 246 (17.6%) | 0.0356 |

[a] 61 respondents did not identify their gender

[b] categories not mutually exclusive

**eFigure 1. Top new or additional concerns related to COVID-19 and physical distancing measures reported by survey respondents, May 2020.**

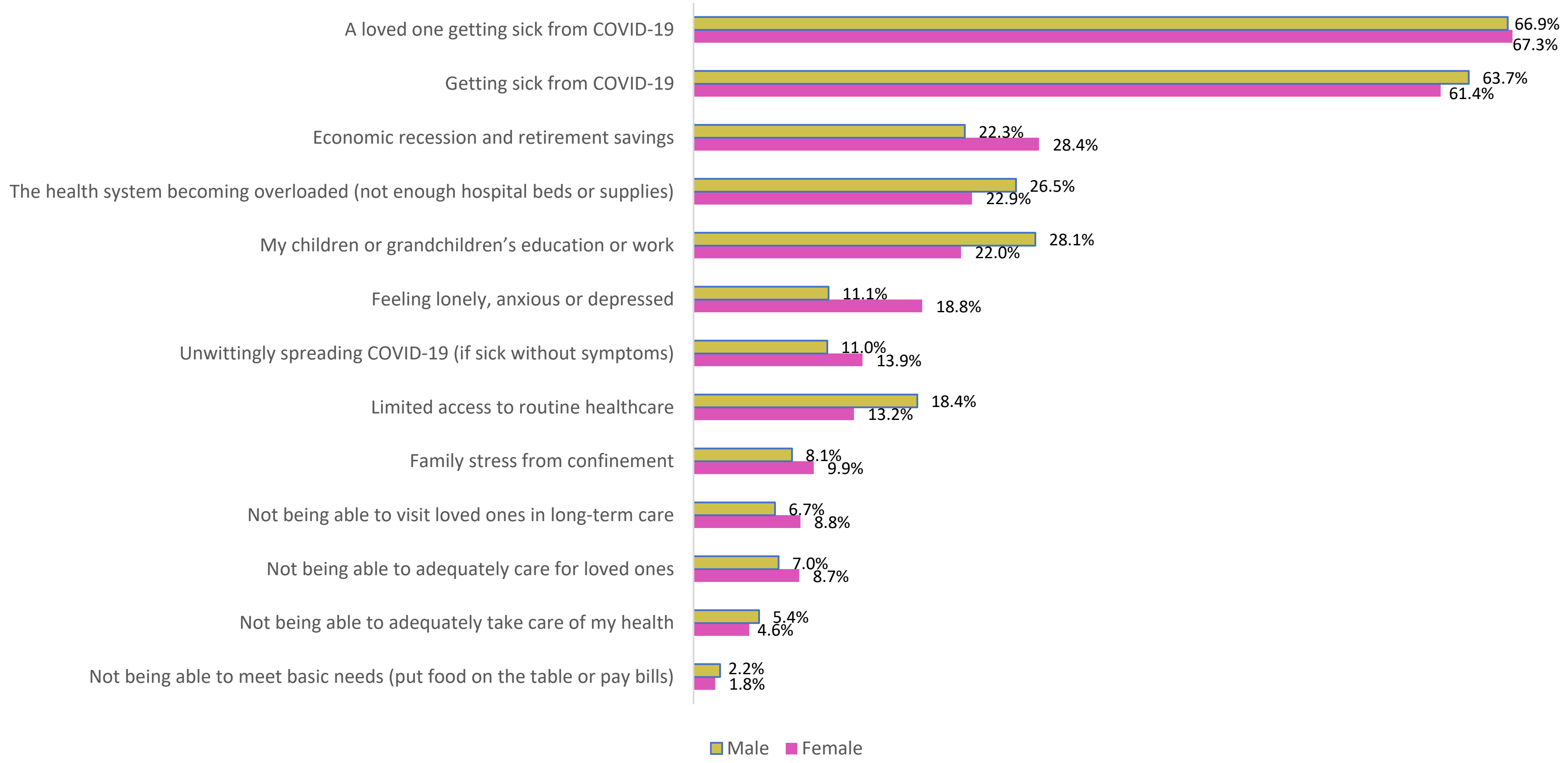
